# A genetically supported drug repurposing pipeline for diabetes treatment using electronic health records

**DOI:** 10.1101/2022.12.14.22283414

**Authors:** Megan M. Shuey, Kyung Min Lee, Jacob Keaton, Nikhil K. Khankari, Joseph H. Breeyear, Venexia M. Walker, Donald R. Miller, Kent R. Heberer, Peter D. Reaven, Shoa L. Clarke, Jennifer Lee, Julie A. Lynch, Marijana Vujkovic, Todd L. Edwards

## Abstract

**Objectives:** The identification of novel uses for existing drug therapies has the potential to provide a rapid, low-cost approach to drug (re)discovery. In the current study we developed and tested a genetically-informed drug-repurposing pipeline for diabetes management.

**Design:** We developed and tested a genetically-informed drug-repurposing pipeline for diabetes management. This approach mapped genetically predicted gene expression signals from the largest genome-wide association study for type 2 diabetes mellitus to drug targets using publicly available databases to identify drug-gene pairs. These drug-gene pairs were then validated using a two-step approach: 1) a self-controlled case-series (SCCS) using electronic health records from a discovery and replication population, and 2) Mendelian randomization (MR).

**Setting:** The SCCS experiments were completed using two EHRs: the Million Veterans Program (USA) as the discovery and the Vanderbilt University Medical Center (Tennessee, USA) as the replication.

**Results:** After filtering on sample size, 20 candidate drug-gene pairs were validated and various medications demonstrated evidence of glycemic regulation including two anti-hypertensive classes: angiotensin-converting enzyme inhibitors as well as calcium channel blockers (CCBs). The CCBs demonstrated the strongest evidence of glycemic reduction in both validation approaches (SCCS HbA1c and glucose reduction: -0.11%, p=0.01 and -0.85 mg/dL, p=0.02, respectively; MR: OR=0.84, 95% CI=0.81, 0.87, p=5.0×10-25).

**Conclusions:** Our results support CCBs as a strong candidate medication for blood glucose reduction in addition to cardiovascular disease reduction. Further, these results support the adaptation of this approach for use in future drug-repurposing efforts for other conditions.

**Summary Boxes:** *Section 1: What is already known on this topic:* Medications with genetic support are significantly more likely to make it through clinical trials. Section 2: What this study adds
Our results identified two anti-hypertensive medication classes, calcium channel blockers and angiotensin-converting enzyme inhibitors, as genetically supported drug-repurposing targets that demonstrated glycemic measurement reduction in real-world clinical populations. These results suggest patients with diabetes or pre-diabetes could benefit from preferential use of these medication classes when they present with comorbid hypertension or other cardiovascular conditions. Finally, this study demonstrates a successful implementation of a novel genetically-supported drug-repurposing pipeline for diabetes treatment that can be readily adapted and applied to other diseases and as such it has the potential to identify/prioritize drug repurposing targets for these other conditions.

## Introduction

An estimated 463 million individuals globally have a diabetes diagnosis^1^ and by 2045 this number is expected to reach 700 million. These rising case numbers are primarily due to type 2 diabetes (T2D) cases, the most prevalent form of the disease^1^. Major comorbidities of T2D include heart disease, peripheral artery disease, stroke, eye disease, kidney disease, and neuropathy^2-4^.

Early stages of glycemic dysregulation can be effectively managed with behavioral modifications and metformin monotherapy. However, as T2D progresses, the concurrent use of multiple medications acting on distinct pathophysiologic pathways is often needed to control blood glucose levels and reduce development of complications as T2D progresses^5-8^. Regardless, a high proportion of patients with T2D demonstrate poor or inadequate glycemic control^9^.

The reason for poor glycemic control is multifactorial. These may include poor medication adherence due to undesired side-effects and costs^10,11^, reduced treatment efficacy^12,13^, and ineffective management of secondary metabolic and glucoregulatory dysfunction^14,15^. Efforts to improve glycemic control include outreach to improve understanding of disease and development of new medications that target glucoregulatory dysfunctions to slow disease progression^16-18^. T2D and comorbidities are also often treated separately, which may lead to increased morbidity and mortality due to polypharmacy^19^.

Drug repurposing provides an efficient, cost-effective means to increase therapeutic options by identifying medications that are currently approved for other indications for treatment of a disease^20^. An example of this is acetylsalicylic acid, marketed as the analgesic Aspirin in 1899, which was subsequently discovered to both inhibit platelet aggregation^21^ and lower glucose^22^. The advance of high-throughput technologies, such as genomics and transcriptomics, supports the development of new computational approaches for drug repurposing and identification of possible adverse drug events^23^. These techniques leverage the highly druggable nature of human disease genes^24^ which may be implicated in diseases other than those indicated for derived medications^25-27^.

Serendipitous discoveries, usually a consequence of clinical observations of patients being treated for other conditions^28^, have historically driven drug repurposing. Findings from real-world observations, however, are subject to biases, such as confounding by indication, reverse causality, and selective data missingness^29^. Mendelian randomization (MR) may overcome some of the limitations of observational epidemiology^29^. Here we propose a computational drug repurposing approach that identifies potential therapeutic candidates for T2D by 1) applying computational methods to genome-wide association study (GWAS) results, 2) evaluating these candidates through a observational self-controlled case series conducted in the electronic health records (EHR) using serendipitous clinical observations in the Department of Veterans Affairs (VA) Corporate Data Warehouse and Vanderbilt University Medical Center’s synthetic derivative (VUMC SD), and 3) proxying the candidate’s drug exposure as predicted expression of the therapeutic gene target using S-PrediXcan in a MR analysis.

## Methods

### Gene-based medication discovery

As described previously^30^, we used summary statistics from the largest multi-ethnic GWAS of T2D, which included over 1.4 million participants, to estimate genetically predicted gene expression (GPGE) of individual genes and performed a transcriptome-wide association study (TWAS) using s-PrediXcan. The imputation of GPGE was based on version 7 predictors for 52 tissues including 48 from GTEx, two kidney tissues (glomerulus and tubule)^31^, and two tissues from an alpha and beta islet cell reference^32^ and signals were refined using colocalization analyses.^30^ This analysis identified 695 significant genes that we mapped to multiple drug targets using publicly available databases: ChEMBL^33^, Drug Gene Interaction Database (DGIdb)^34^, and National Cancer Institute Drug Dictionary^35^. The mapped genes were further refined by pairing the direction of GPGE and T2D risk with drug effects. For example, genes with increased GPGE that were associated with a decrease in T2D risk would need to map to a drug that acts as an activator or agonist of the associated protein or pathway.

Conversely, drugs that act as an antagonist, inhibitor or blocker would correspond with a gene for which increasing GPGE associated with an increase in T2D risk. This process is visualized in Figure 1, steps 1-4. Drug-gene pairs with correlated functions that were not identified previously for use in diabetes management were then included as experimental medications for the subsequent analyses to evaluate their potential for repurposing including a self-controlled case series and MR studies.

**Figure 1.**
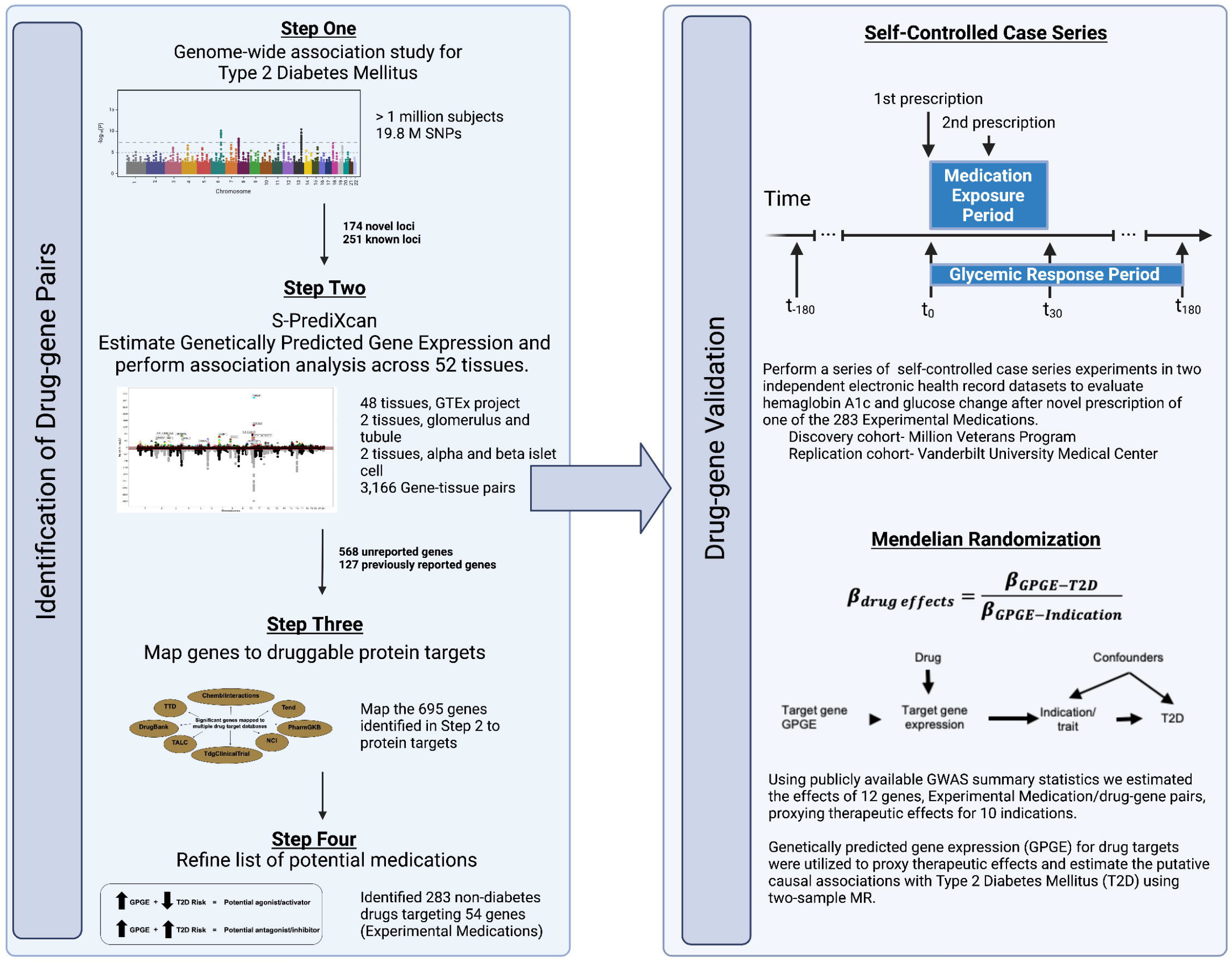
Study design. Steps 1-4 describe the process of identifying novel drug targets based on genetic evidence using a previously published genome-wide association study for Type 2 Diabetes Mellitus. Following identification of potential medications based on drug-gene pairing the results are validated using two approaches, self-controlled case series and mendelian randomization (MR). The general design of these two approaches are described briefly.

### Self-controlled case series data sources

Two separate EHR systems were identified as data sources for the self-controlled case series. The discovery population included clinical, prescription, and laboratory data from the Corporate Data Warehouse (CDW) of the Veterans Health Administration (VHA), a national US data repository that provides access to the EHRs of all individuals who received care in the VHA. All study variables were extracted from the CDW in April 2020 by an experienced programmer on Microsoft SQL Server via the VA Informatics and Computing Infrastructure (VINCI) computing environment^36^.

The replication site data were collected from the VUMC SD, a de-identified copy of the electronic medical record with Health Insurance Portability and Accountability Act of 1996 (HIPAA) identifiers removed^37^. The SD contains clinical data on approximately 3.2 million individuals and includes basic demographics; text from clinical care notes; laboratory values; inpatient and outpatient medication data; International Classification of Disease (ICD) and Current Procedural Terminology (CPT) codes; and other diagnostic reports. Drug exposures were identified using previously described electronic-prescribing tools and MedEx^38^. The utility of these methods for medication extraction from the EHR have been shown previously^39,40^. All valid medication exposures required one of the following indications to be documented: route, frequency, dose, or duration. All study variables were extracted from the SD by an experienced programmer in January 2020.

### Self-controlled case series study design

We utilized a self-controlled case series study design within the discovery and replication populations to evaluate the effects of novel medication use on hemoglobin A1c (HbA1c) and glucose. The design varied slightly between the two sites to accommodate variations in data availability and population. These variances are discussed below. The general design of this study is displayed in Figure 2.

**Figure 2.**
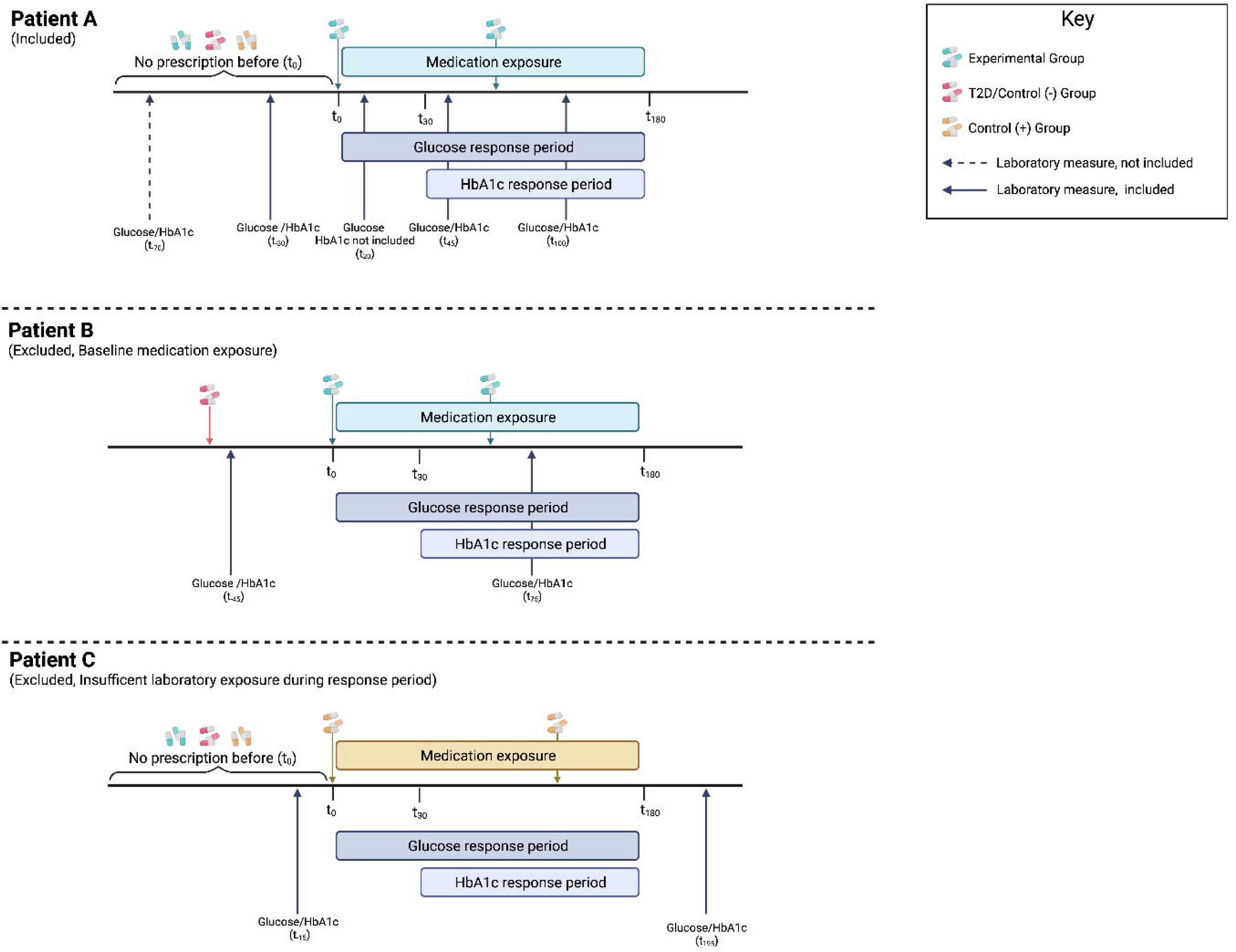
Examples of the medication exposure time period for the self-controlled case series for determination of patient inclusion or exclusion. Patient A demonstrates a patient that would be included in the study based on study design. Specifically, this patient was prescribed a medication belonging to the experimental group and had a subsequent mention of the medication in their records in the following 6 months. They had no documented exposure to a medication belonging to one of the other medication groups in the period preceding t_0_. They also had glucose and hemoglobin A1c (HbA1c) measures collected in the six months before medication exposure and during the response periods. Conversely, patients B and C were excluded from this study. Patient B represents patients that were excluded from the study because they had a medication exposure from another medication group prior to t_0_. Patient C is an example of a patient that would be excluded due to insufficient data. In this example, Patient C had no glucose and HbA1c measurements in the six-month exposure period. Likewise, patients without a glucose or HbA1c in the six months before t_0_ would also be excluded from the study due to insufficient laboratory exposure.

Medications evaluated in this series were grouped in to three sets: 1) experimental, the gene-based medication set described previously; 2) diabetes/control (−), medications that are prescribed for the treatment of T2DM; and 3) control (+), medications belonging to classes that had previously described increasing effects on glucose or HbA1c or were implicated in a previous MR study^41^. The complete list of medications and their corresponding set are available in Supplemental Tables 1a and b. To ensure the effects on glucose or HbA1c were due to a specific medication, each individual medication series included patients that were prescribed the specific medication when the drug was first prescribed, t_0_, but excluded patients prescribed a medication in one of the other groups ever before, simultaneously at t_0_, or in the six-month follow-up period.

Further, in the discovery set the VA patients were only included in a medication series if they received at least one subsequent refill of the given medication within 180 days of t_0_, and had at least 90 days of cumulative exposure. Because records of a patient’s prescription fill is less readily available in the VUMC SD, e.g. many patients will fill their prescriptions at a non-VUMC pharmacy so records of prescription pick-ups are absent in their record, this study required included subject EHRs in VUMC SD to have at least two mentions of the medication within 180 days of the initial drug mention, t_0_, to proxy prescription refill.

Finally, all patients included in the medication series were required to have both a baseline and follow-up HbA1c and glucose measured to evaluate response. HbA1c and glucose measures were restricted to outpatient measures and non-physiologic measures were excluded, e.g. HbA1c less than 3% or greater than 18% and glucose less than 5 mg/dL or greater than 2,750 mg/dL. In the replication population we also excluded all laboratory measures that were collected within 9 months of a pregnancy ICD code or laboratory test (Supplemental Table 2). Because the discovery cohort is >90% male this exclusion was not applied.

The change in laboratory measure was defined as absolute change. The absolute change was calculated by taking the difference between the follow-up measure and the baseline measure. Baseline HbA1c was defined as the most recent measure in the six months prior to drug initiation. Follow-up HbA1c measure was defined as the first measure after 30 days and within the six months following drug initiation (Figure 2). For glucose, we used the mean of all measures in the six months prior to drug initiation as the baseline value and the mean of all measures in the six months after drug initiation as the follow-up value.

### Statistical Analysis

We assessed patient characteristics at drug initiation by comparing demographics, smoking status, body mass index, glycemic status, and Charlson Comorbidity Index categories across the three drug groups for both study populations. Using a self-controlled case series design, we performed pairwise t-test to determine statistical significance of the change in laboratory measures. Drugs with a sample size less than 30 in the discovery population were not analyzed to increase precision. We used SAS 9.2 (Cary, NC) for all data preparation and analysis in the discovery population and Rv3.2 for the replication population. We also performed a random-effects meta-analysis to estimate between-study and within medication class effect sizes for glucose and HbA1c change using STATA.

### Mendelian randomization

The MR analysis was structured around a subset of 12 genes identified as drug-gene pairs in Figure 1, step 4. Two-sample MR was conducted by leveraging summary statistics from existing GWAS on ten different drug indications (angina^42^, atrial fibrillation^43^, bipolar disorder^44^, coronary artery disease^45^, congestive heart failure^46^, epilepsy^47^, glaucoma^48^, pain^49^, rheumatoid arthritis^50^, systolic blood pressure^51^) in tandem with summary statistics from the largest T2D GWAS to date^30^. For the 12 genes, the MR analysis utilized only tissues with statistically significant GPGE (P<0.05) as the instrument and was conducted via the “TwoSampleMR” R package^52^.

We replicated a previously published approach using S-PrediXcan summary statistics as the instrumental variable in an MR analysis to proxy therapeutic targets^41^.Tissue-specific GPGE summary statistics for each trait and T2D were combined to calculate the IVW MR association as follows:

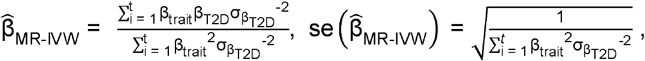

where β_trait_represents GPGE per trait; and β_T2D_ and 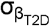 represent T2D GPGE and standard error, for *t* number of statistically significant GPGE tissues, respectively.

Corresponding odds ratios (ORs) and 95% confidence intervals (95% CI) were calculated using 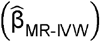 and se 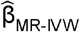.

The estimates (and 95% CIs) obtained from the MR analysis represent T2D risk per standard deviation change in gene expression. Furthermore, MR Egger regression was used to evaluate directional pleiotropy^53^. Multivariable MR (MVMR) was conducted to estimate adjusted MR effects for indications that were correlated and shared therapeutic gene targets (e.g., ACE inhibitors are used to treat hypertension and congestive heart failure)^54^.

### Patient and Public Involvement

For the VA discovery population, patient clinical data were analyzed as part of the Leveraging Electronic Health Information to Advance Precision Medicine research protocol, which has been approved by institutional review boards and research committees at 3 VA Medical Centers (Salt Lake City, Palo Alto, and West Haven) with approved waivers of informed consent and HIPAA authorization. For the replication population, VUMC SD, the associated project received non-human subjects determination and approval from the VUMC Institutional Review Board.

## Results

### Computational drug repurposing approach

We used a multi-step computational approach to drug repurposing that leveraged large scale GWAS and EHR data to identify and test potential non-diabetes medications for use in hemoglobin (HbA1c) and glucose control (Figure 1). Steps 1-4 describe the process of identifying drug-gene pairs for evaluation and validation. The evaluation stage takes a two-pronged approach to evaluate these drug-gene pairs: a) a self-controlled case series to evaluate the response of HbA1c and glucose to novel medication exposure and b) a Mendelian randomization study to proxy therapeutic effects of a subset of the identified gene-drug pairs.

### Gene-based medication discovery (Figure 1, steps 1-4)

Briefly, genes were identified in a previous large-scale GWAS and TWAS of diabetes risk^30^. Publicly available databases of drug gene targets, indications, and interactions were consulted to identify drug-gene pairs. Drugs were selected such that the drug targeted a gene that was associated with diabetes risk via GPGE, the drug was not used in diabetes management, and where the action of the drug would be predicted to mitigate increases in diabetes risk.

Summary statistics for 19.8 million single nucleotide polymorphisms were utilized in S-PrediXcan models for GPGE estimation across 52 tissues. We identified 695 unique genes that were associated with diabetes risk at a p-value threshold of 1.92×10^−7^, including: 568 genes for which a relationship was not previously reported and 127 with a known relationship to diabetes. We further refined the list of target drug-gene pairs by comparing medication and gene effects. For example, if a drug was an activator or agonist of the associated pathway or enzyme activity, we expect that increasing GPGE of the corresponding gene would be associated with decreased diabetes risk, and vice-versa for inhibitors. We identified 283 drugs with repurposing potential for diabetes that targeted 54 genes, 7.7% of the unique genes reported from the TWAS.

### Self-controlled case series

To test for the impact of novel medication start on HbA1c and glucose using EHRs, we designed a self-controlled case series to evaluate the change in these laboratory measures in the six months following medication initiation (Figure 2). We restricted medication starts to a single experimental medication, defined by a gene-drug pair identified previously, with no preceding prescription of control medications (established medications known to raise or lower HbA1c and glucose). These medications were classified as glucose-reducing or glucose-increasing control medications based on prior knowledge of their effects on HbA1c or glucose. Overall, 68 medications were identified as possible control medications (Supplemental Table 1a and b). Further, we repeated this self-controlled case series for the glucose-reducing or glucose-increasing medications using identical restrictions on medication exposure and novel start to evaluate design performance (Figure 2). We performed the self-controlled case series independently in two large EHR systems, the Veteran’s Administration (VA) and Vanderbilt University Medical Center Synthetic Derivative (VUMC SD), separately for HbA1c and glucose.

#### Hemoglobin A1c

The VA discovery population for the HbA1c self-controlled case series included 124,357 patients: 40,780 (32.8%) in the T2D/glucose-reducing group, 25,170 (20.2%) in the glucose-increasing group, and 58,407 (47.0%) in the experimental group. The baseline characteristics of these patients are summarized in Supplemental Table 3a. The VUMC SD replication population was one-tenth the size of the VA population, including 15,365 patients: 4,439 (28.9%) in the T2D/glucose-reducing group, 3,912 (25.5%) in the glucose-increasing group, and 7,014 (45.6%) in the experimental group. The baseline characteristics of these patients are summarized in Supplemental Table 3b.

The paired t-test results for HbA1c change in both the discovery and replication populations are summarized in Table 1. Analyzed medications were required to have at least 30 prescriptions in both sites. All T2D/glucose-reducing medications demonstrated substantial reductions in HbA1c in the discovery population with similar results in the replication set. The greatest reduction in HbA1c in the VA population was glyburide and glipizide, second-generation oral sulfonylureas, with mean changes of - 1.3% and -1.1%, respectively (p<0.001). The control (+) medications had more inconsistent effects on HbA1c across the two sites. Of the 16 glucose-increasing medications, only amitriptyline had evidence to support an increase in HbA1c in both populations. Seven additional medications were associated with increases in the discovery population but not in the replication. We observed consistent direction of efforts with these 7 medications in the replication population but the substantially lower sample sizes potentially reduced the power to detect effects.

**Table 1.** HbA1c paired t-test results from VA LEAPS with replication at Vanderbilt (N>=30 at both sites)

|  |  | VA LEAPS Overall |  |  |  |  | Vanderbilt Overall |  |  |  |  |
| --- | --- | --- | --- | --- | --- | --- | --- | --- | --- | --- | --- |
|  |  | 95% C.I. |  |  |  |  | 95% C.I. |  |  |  |  |
| GROUP | DRUG | n | Mean Δ | Lower | Upper | p | n | Mean Δ | Lower | Upper | p |
| Glucose decreasing | GLIPIZIDE | 5289 | -1.0629 | -1.1142 | -1.0117 | <2.2E16 | 429 | -0.6014 | -0.7673 | -0.4355 | 4.39E-12 |
| Glucose decreasing | GLYBURIDE | 6428 | -1.3495 | -1.4023 | -1.2968 | <2.2E16 | 225 | -0.7409 | -0.9786 | -0.5032 | 3.67E-09 |
| Glucose decreasing | INSULIN | 6695 | -0.7686 | -0.8156 | -0.7216 | <2.2E16 | 1558 | -0.7970 | -0.8950 | -0.6991 | <2.2E16 |
| Glucose decreasing | METFORMIN | 20954 | -0.9659 | -0.9888 | -0.9429 | <2.2E16 | 1732 | -0.7761 | -0.8561 | -0.6961 | <2.2E16 |
| Glucose decreasing | PIOGLITAZONE | 300 | -0.3711 | -0.5031 | -0.2391 | 6.29E-08 | 30 | -0.2400 | -0.5396 | 0.0596 | 0.1122 |
| Glucose decreasing | LIRAGLUTIDE | 72 | -0.7554 | -1.0457 | -0.4651 | 5.16E-07 | 34 | -0.4265 | -0.8960 | 0.0430 | 7.36E-02 |
| Glucose decreasing | GLIMEPIRIDE | 74 | -0.6734 | -0.9896 | -0.3572 | 3.57E-05 | 302 | -0.7007 | -0.8781 | -0.5232 | 1.23E-13 |
| Glucose decreasing | SITAGLIPTIN | 54 | -0.2945 | -0.5816 | -0.00733 | 0.0446 | 35 | -0.4257 | -0.9285 | 0.0770 | 0.0944 |
| Experimental | LISINOPRIL | 13265 | -0.0326 | -0.0446 | -0.0207 | 1.48E-07 | 1145 | -0.2383 | -0.3156 | -0.1610 | 1.96E-09 |
| Experimental | BENAZEPRIL | 408 | -0.0875 | -0.1443 | -0.0307 | 0.0026 | 68 | -0.3191 | -0.6207 | -0.0176 | 0.0384 |
| Experimental | VERAPAMIL | 310 | -0.0937 | -0.1691 | -0.0183 | 0.0150 | 81 | -0.2185 | -0.4355 | -0.0016 | 0.0484 |
| Experimental | PRAVASTATIN | 2837 | -0.023 | -0.0432 | -0.00281 | 0.0256 | 284 | -0.2517 | -0.4062 | -0.0971 | 0.0015 |
| Experimental | ENALAPRIL | 379 | -0.0527 | -0.1021 | -0.00317 | 0.0371 | 123 | 0.0691 | -0.1326 | 0.2708 | 0.4989 |
| Experimental | ATORVASTATIN | 6994 | 0.0135 | -0.00316 | 0.0301 | 0.1123 | 619 | -0.1924 | -0.2967 | -0.0881 | 0.0003 |
| Experimental | DICLOFENAC | 1585 | -0.0205 | -0.0458 | 0.00486 | 0.1132 | 238 | -0.0445 | -0.1724 | 0.0833 | 0.4933 |
| Experimental | LOVASTATIN | 1336 | -0.017 | -0.0468 | 0.0129 | 0.2652 | 172 | -0.2291 | -0.3885 | -0.0697 | 0.0051 |
| Experimental | SILDENAFIL | 6198 | -0.00746 | -0.0225 | 0.00762 | 0.3324 | 51 | -0.6882 | -1.2262 | -0.1503 | 0.0132 |
| Experimental | LIDOCAINE | 242 | 0.0183 | -0.035 | 0.0716 | 0.4985 | 567 | -0.1074 | -0.1924 | -0.0224 | 0.0134 |
| Experimental | ISOSORBIDE DINITRATE | 58 | 0.0737 | -0.1642 | 0.3117 | 0.5373 | 177 | -0.0130 | -0.1621 | 0.1361 | 0.8637 |
| Experimental | RAMIPRIL | 59 | 0.022 | -0.1313 | 0.1752 | 0.7750 | 96 | -0.1469 | -0.3592 | 0.0654 | 0.1729 |
| Experimental | ISOSORBIDE | 122 | -0.0198 | -0.1584 | 0.1188 | 0.7775 | 171 | -0.0117 | -0.1658 | 0.1424 | 0.8811 |
| Experimental | SIMVASTATIN | 16840 | -0.00131 | -0.0107 | 0.00806 | 0.7839 | 561 | -0.1422 | -0.2845 | -0.0342 | 0.0099 |
| Experimental | LEVETIRACETAM | 132 | -0.0118 | -0.1091 | 0.0856 | 0.8116 | 75 | -0.0267 | -0.3174 | 0.2641 | 0.8555 |
| Experimental | PREGABALIN | 221 | 0.0132 | -0.0964 | 0.1229 | 0.8122 | 36 | -0.0528 | -0.4504 | 0.3448 | 0.7891 |
| Experimental | LAMOTRIGINE | 290 | -0.00783 | -0.0813 | 0.0657 | 0.8341 | 46 | -0.2739 | -0.7942 | 0.2464 | 0.2947 |
| Experimental | ROSUVASTATIN | 1317 | -0.00338 | -0.0417 | 0.0349 | 0.8625 | 285 | -0.0874 | -0.2068 | 0.0320 | 0.1509 |
| Experimental | ASPIRIN | 2398 | -0.00228 | -0.0327 | 0.0281 | 0.8832 | 1834 | -0.1677 | -0.2182 | -0.1171 | 1.01E-10 |
| Experimental | TOPIRAMATE | 563 | 0.00081 | -0.0369 | 0.0386 | 0.9664 | 53 | -0.3113 | -0.7096 | 0.0870 | 0.1228 |
| Glucose increasing | HYDROCHLOROTHIAZIDE | 4972 | 0.0791 | 0.0609 | 0.0972 | < 2.2E-16 | 664 | 0.0541 | -0.0188 | 0.1269 | 0.1457 |
| Glucose increasing | PREDNISONE | 604 | 0.2436 | 0.166 | 0.3212 | 1.76E-09 | 412 | 0.0080 | -0.1029 | 0.1189 | 0.8871 |
| Glucose increasing | PROPRANOLOL | 785 | 0.1198 | 0.0648 | 0.1747 | 2.35E-05 | 94 | 0.0989 | -0.0826 | 0.2805 | 0.2820 |
| Glucose increasing | FLUOXETINE | 1753 | -0.0356 | -0.0631 | -0.00806 | 0.0113 | 176 | 0.0097 | -0.1351 | 0.1544 | 0.8954 |
| Glucose increasing | PAROXETINE | 560 | 0.0639 | 0.00905 | 0.1188 | 0.0225 | 55 | -0.0091 | -0.2567 | 0.2385 | 0.9416 |
| Glucose increasing | AMITRIPTYLINE | 886 | 0.0526 | 0.00413 | 0.1011 | 0.0335 | 141 | 0.1816 | 0.0268 | 0.3364 | 0.0219 |
| Glucose increasing | NORTRIPTYLINE | 342 | 0.0617 | 0.00315 | 0.1202 | 0.0390 | 32 | 0.0469 | -0.2187 | 0.3124 | 0.7213 |
| Glucose increasing | METOPROLOL | 2535 | 0.0261 | 0.00111 | 0.051 | 0.0406 | 520 | -0.0223 | -0.1315 | 0.0868 | 0.6882 |
| Glucose increasing | CITALOPRAM | 2838 | -0.0217 | -0.0427 | -0.00068 | 0.0431 | 339 | -0.0071 | -0.1372 | 0.1230 | 0.9148 |
| Glucose increasing | ATENOLOL | 1906 | 0.0214 | -0.00719 | 0.0501 | 0.1421 | 240 | 0.1521 | -0.0068 | 0.3109 | 0.0605 |
| Glucose increasing | CARVEDILOL | 481 | 0.0423 | -0.0294 | 0.114 | 0.2469 | 247 | 0.1810 | 0.0079 | 0.3540 | 0.0405 |
| Glucose increasing | OLANZAPINE | 185 | 0.0487 | -0.0344 | 0.1319 | 0.2492 | 54 | 0.0111 | -0.1708 | 0.1931 | 0.9030 |
| Glucose increasing | SERTRALINE | 4407 | -0.0103 | -0.0282 | 0.0075 | 0.2561 | 151 | -0.1265 | -0.2959 | 0.0430 | 0.1423 |
| Glucose increasing | RISPERIDONE | 424 | 0.0343 | -0.0273 | 0.096 | 0.2742 | 47 | -0.3809 | -0.8242 | 0.0625 | 0.0905 |
| Glucose increasing | QUETIAPINE | 766 | 0.0161 | -0.014 | 0.0462 | 0.2927 | 36 | 0.0222 | -0.2910 | 0.3355 | 0.8863 |
| Glucose increasing | EZETIMIBE | 102 | -0.0309 | -0.1221 | 0.0603 | 0.5031 | 80 | 0.1925 | -0.1370 | 0.5220 | 0.2484 |

Of the initial 283 experimental medications identified by drug-gene pairs, only 20 (7.1%) had sufficient data for inclusion in the HbA1c self-controlled case series. This set included four angiotensin-converting enzyme (ACE) inhibitors. Two demonstrated substantial reductions in HbA1c in the discovery and replication populations, and one, enalapril, had evidence for a reduction in the discovery but not the replication set.

Ramipril did not consistently reduce HbA1c in either set. Lisinopril demonstrated the largest reduction (p<0.001, mean change VA= -0.03% and VUMC=-0.24%) in both populations. The calcium channel blocker verapamil also demonstrated reductions in HbA1c in both sets. Most statins had inconsistent effects. Pravastatin demonstrated a decrease in HbA1c in both populations.

#### Glucose

The VA discovery population for the glucose self-controlled case series included 678,501 patients: 51,645 (7.6%) in the T2D/glucose-reducing group, 235,493 (34.7%) in the glucose-increasing group, and 391,363 (57.7%) in the experimental group. The baseline characteristics of these patients are summarized in Supplemental Table 4a. The VUMC SD replication population was about one-tenth of the size of the VA population, including 67,155 patients: 11,889 (17.7%) in the T2D/glucose-reducing group, 9,371 (14.0%) in the glucose-increasing group, and 45,895 (68.3%) in the experimental group. The baseline characteristics of these patients are summarized in Supplemental Table 4b.

The paired t-test results for glucose change in both the discovery and replication populations are summarized in Table 2. Consistent with the HbA1c results, glyburide and glipizide had the most substantial decrease in glucose (p<0.001) in the discovery set. For the most part, all T2D/glucose-reducing medications demonstrated decreases in glucose. Some medications, such as sitagliptin, decreased glucose in the replication but not the discovery set (VA, p=0.44 and VUMC, p=0.002). These results suggest high variability in random glucose measurements may decrease statistical precision. The glucose-increasing medications demonstrated similar variability in results with dexamethasone exhibiting the only consistent increase in glucose across both populations (p<0.001).

**Table 2.** Glucose paired t-test results from VA LEAPS with replication at Vanderbilt (N>=30 at both sites)

|  |  | VA LEAPS Overall |  |  |  |  | Vanderbilt Overall |  |  |  |  |
| --- | --- | --- | --- | --- | --- | --- | --- | --- | --- | --- | --- |
|  |  | 95% C.I. |  |  |  |  | 95% C.I. |  |  |  |  |
| GROUP | DRUG | n | Mean Δ | Lower | Upper | p | n | Mean Δ | Lower | Upper | p |
| Glucose decreasing | GLYBURIDE | 9711 | -57.4723 | -59.4317 | -55.5128 | < 2.2E-16 | 651 | -6.4471 | -15.0083 | 2.1141 | 0.1397 |
| Glucose decreasing | GLIPIZIDE | 7025 | -41.2591 | -43.2906 | -39.2276 | < 2.2E-16 | 1117 | -9.7684 | -15.7292 | 3.8076 | 0.0013 |
| Glucose decreasing | METFORMIN | 25046 | -34.2185 | -35.1366 | -33.3004 | < 2.2E-16 | 4381 | -14.9646 | -17.6653 | -12.2639 | < 2.2E-16 |
| Glucose decreasing | INSULIN | 8004 | -29.6005 | -32.062 | -27.1391 | < 2.2E-16 | 4575 | -5.4490 | -8.8399 | -2.0581 | 0.0016 |
| Glucose decreasing | LIRAGLUTIDE | 73 | -28.4548 | -45.8076 | -11.1021 | 0.0001 | 52 | -12.4888 | -37.1523 | 12.1747 | 0.3142 |
| Glucose decreasing | PIOGLITAZONE | 345 | -9.2861 | -15.155 | -3.4173 | 0.0020 | 100 | -9.5621 | -26.8865 | 7.7624 | 0.2761 |
| Glucose decreasing | GLIMEPIRIDE | 81 | -20.9352 | -37.9492 | -3.9211 | 0.0165 | 703 | -17.8257 | -25.9716 | -9.6797 | 1.98E-05 |
| Glucose decreasing | SITAGLIPTIN | 54 | -6.3206 | -22.8982 | 10.257 | 0.4478 | 77 | -26.3620 | -42.8639 | -9.8602 | 0.0021 |
| Experimental | SIMVASTATIN | 127647 | 0.6144 | 0.5117 | 0.7172 | < 2.2E-16 | 3498 | 0.7349 | -1.8133 | 3.2831 | 0.5718 |
| Experimental | ATORVASTATIN | 22491 | 1.0187 | 0.7367 | 1.3006 | 5.40E-12 | 2829 | -1.4375 | -4.5069 | 1.6318 | 0.3585 |
| Experimental | SILDENAFIL | 32811 | 0.5618 | 0.3186 | 0.8049 | 7.67E-06 | 461 | 6.2037 | -1.8846 | 14.2919 | 0.1324 |
| Experimental | PRAVASTATIN | 12505 | 0.7337 | 0.4069 | 1.0605 | 1.35E-05 | 1660 | 3.4621 | -0.7323 | 7.6565 | 0.1056 |
| Experimental | TOPIRAMATE | 3612 | 0.9559 | 0.3895 | 1.5222 | 0.0009 | 405 | 1.4338 | -4.6082 | 7.4758 | 0.6411 |
| Experimental | ROSUVASTATIN | 4800 | 1.0157 | 0.3969 | 1.6345 | 0.0013 | 1411 | 2.5107 | -1.1692 | 6.1906 | 0.1810 |
| Experimental | LOVASTATIN | 12800 | 0.3971 | 0.0895 | 0.7048 | 0.0114 | 770 | 3.9668 | -1.0250 | 8.9586 | 0.1192 |
| Experimental | VERAPAMIL | 3712 | -0.819 | -1.5727 | -0.0654 | 0.0332 | 715 | -2.0611 | -6.4948 | 2.3726 | 0.3617 |
| Experimental | OXCARBAZEPINE | 266 | 2.6131 | 0.1343 | 5.0919 | 0.0389 | 376 | -12.8438 | -22.9367 | -2.7508 | 0.0128 |
| Experimental | FLUVASTATIN | 1475 | 0.8816 | 0.0237 | 1.7394 | 0.0440 | 39 | 1.0832 | -14.0903 | 16.2566 | 0.8859 |
| Experimental | ENALAPRIL | 2423 | -0.7362 | -1.5228 | 0.0505 | 0.0666 | 922 | 1.3151 | -2.5047 | 5.1348 | 0.4994 |
| Experimental | LAMOTRIGINE | 2148 | 0.7441 | -0.0665 | 1.5547 | 0.0720 | 467 | 2.7112 | -3.8419 | 9.2643 | 0.4166 |
| Experimental | DICLOFENAC | 9858 | 0.3563 | -0.0532 | 0.7658 | 0.0882 | 1328 | 0.4467 | -3.4450 | 4.3384 | 0.8219 |
| Experimental | PREGABALIN | 996 | 1.1986 | -0.1859 | 2.583 | 0.0896 | 400 | -4.5174 | -13.3906 | 4.3558 | 0.3175 |
| Experimental | LISINOPRIL | 97013 | 0.083 | -0.0606 | 0.2266 | 0.2574 | 6616 | 1.0676 | -0.9022 | 3.0374 | 0.9020 |
| Experimental | SALMON CALCITONIN | 550 | 0.8704 | -0.6668 | 2.4076 | 0.2665 | 72 | -0.4616 | -11.1156 | 10.1925 | 0.9314 |
| Experimental | MINOXIDIL | 154 | 2.1982 | -1.7341 | 6.1304 | 0.2712 | 415 | -1.1851 | -9.7363 | 7.3661 | 0.7855 |
| Experimental | DOFETILIDE | 132 | 2.4776 | -2.1688 | 7.1241 | 0.2934 | 52 | -3.1943 | -17.5331 | 11.1445 | 0.6566 |
| Experimental | DABIGATRAN ETEXILATE | 754 | 0.8961 | -0.8011 | 2.5933 | 0.3003 | 52 | 2.3017 | -9.1225 | 13.7258 | 0.6876 |
| Experimental | SULINDAC | 2358 | 0.4141 | -0.3778 | 1.2061 | 0.3053 | 64 | -0.8107 | -19.3465 | 17.7251 | 0.9306 |
| Experimental | PROPAFENONE | 220 | 1.9354 | -1.8129 | 5.6837 | 0.3100 | 109 | -2.9841 | -11.3359 | 5.3678 | 0.4803 |
| Experimental | PRILOCAINE | 237 | -1.4097 | -4.3919 | 1.5726 | 0.3527 | 746 | 1.9719 | -2.2340 | 6.1778 | 0.3577 |
| Experimental | ASPIRIN | 23073 | -0.1137 | -0.4183 | 0.191 | 0.4645 | 11303 | -0.3834 | -0.9497 | 1.7165 | 0.5730 |
| Experimental | LEVETIRACETAM | 1126 | 0.4293 | -0.7271 | 1.5856 | 0.4665 | 947 | -4.8177 | -10.3211 | 0.6856 | 0.0861 |
| Experimental | LIDOCAINE | 1560 | 0.363 | -0.7034 | 1.4294 | 0.5044 | 6143 | 1.9847 | 0.1400 | 3.8295 | 0.0350 |
| Experimental | DIPYRIDAMOLE | 753 | 0.4788 | -1.1379 | 2.0955 | 0.5611 | 70 | 5.1318 | -3.2526 | 13.5163 | 0.2262 |
| Experimental | BENZAEPRIIL | 2431 | -0.2343 | -1.0657 | 0.5972 | 0.5806 | 368 | 1.3187 | -5.1389 | 7.7763 | 0.6882 |
| Experimental | RAMIPRIIL | 424 | 0.4899 | -1.7544 | 2.7343 | 0.6681 | 482 | -1.0035 | -6.8555 | 4.8485 | 0.7363 |
| Experimental | ISOSORBIDE | 641 | 0.4894 | -1.7615 | 2.7404 | 0.6695 | 1250 | 0.5219 | -3.9577 | 5.0015 | 0.8192 |
| Experimental | PHENYTOIN | 2529 | 0.1789 | -0.8601 | 1.2178 | 0.7357 | 254 | 2.6984 | -3.7677 | 9.1646 | 0.4119 |
| Experimental | MICONAZOLE | 1551 | -0.1249 | -1.2148 | 0.9651 | 0.8222 | 80 | -6.8415 | -17.0917 | 3.4086 | 0.1878 |
| Experimental | ISOSORBIDE DINITRATE | 963 | -0.1941 | -2.1833 | 1.7951 | 0.8482 | 1256 | -0.2454 | -4.7446 | 4.2539 | 0.9148 |
| Experimental | PRIMIDONE | 720 | -0.0799 | -1.754 | 1.5943 | 0.9254 | 125 | -1.1066 | -11.6441 | 9.4309 | 0.8357 |
| Experimental | FOSINOPRIIL | 10635 | 0.0201 | -0.4368 | 0.477 | 0.9314 | 73 | 1.1213 | -12.3272 | 14.5698 | 0.8685 |
| Experimental | PENTOXIFYLLINE | 571 | 0.0227 | -2.066 | 2.1115 | 0.9829 | 105 | 6.6259 | -12.5793 | 25.8311 | 0.4954 |
| Glucose increasing | DEXAMETHASONE | 616 | 10.787 | 8.2927 | 13.2813 | < 2.2E-16 | 804 | 4.7224 | 1.3065 | 8.1382 | 0.0068 |
| Glucose increasing | HYDROCHLOROTHIAZIDE | 61523 | 2.6293 | 2.464 | 2.7946 | < 2.2E-16 | 1731 | 0.5670 | -1.6252 | 2.7592 | 0.6120 |
| Glucose increasing | PREDNISONE | 8487 | 2.9327 | 2.3714 | 3.494 | < 2.2E-16 | 1058 | 1.3596 | -2.9937 | 5.7130 | 0.5401 |
| Glucose increasing | PROPRANOLOL | 7056 | 1.6425 | 1.0486 | 2.2363 | 1.00E-07 | 171 | 1.1228 | -9.9198 | 12.1654 | 0.8412 |
| Glucose increasing | CARVEDILOL | 2512 | 2.3207 | 1.2185 | 3.4229 | 4.35E-05 | 805 | 4.4634 | -2.0466 | 10.9733 | 0.1787 |
| Glucose increasing | OLANZAPINE | 1930 | 2.7479 | 1.4033 | 4.0925 | 7.14E-05 | 58 | -4.5690 | -13.3503 | 4.2124 | 0.3019 |
| Glucose increasing | AMITRIPTYLINE | 9039 | 0.7909 | 0.2944 | 1.2873 | 0.0018 | 204 | -2.5294 | -10.3204 | 5.2616 | 0.5228 |
| Glucose increasing | ATENOLOL | 23108 | 0.3782 | 0.0884 | 0.668 | 0.0105 | 568 | 1.4609 | -3.0948 | 6.0166 | 0.5290 |
| Glucose increasing | RISPERIDONE | 3493 | 1.01 | 0.2141 | 1.8059 | 0.0129 | 52 | -20.9423 | -55.2099 | 13.3253 | 0.2255 |
| Glucose increasing | METHYLPREDNISOLONE | 193 | 3.4302 | -0.6742 | 7.5346 | 0.1009 | 35 | 20.8881 | 1.4221 | 40.3540 | 0.0005 |
| Glucose increasing | CITALOPRAM | 26808 | -0.1919 | -0.4457 | 0.0618 | 0.1382 | 389 | -3.5900 | -9.4851 | 2.3052 | 0.2319 |
| Glucose increasing | BISOPROLOL | 112 | 3.0967 | -1.0433 | 7.2366 | 0.1411 | 33 | -2.7424 | -20.8768 | 15.3920 | 0.7600 |
| Glucose increasing | PAROXETINE | 5447 | 0.3322 | -0.2405 | 0.9049 | 0.2555 | 108 | 1.6204 | -8.6618 | 11.9026 | 0.7553 |
| Glucose increasing | HYDROCORTISONE | 503 | 1.0903 | -0.8136 | 2.9942 | 0.2611 | 97 | 8.0670 | -0.3042 | 16.4383 | 0.0588 |
| Glucose increasing | METOPROLOL | 19954 | 0.1791 | -0.1492 | 0.5073 | 0.2849 | 1726 | 4.2568 | 1.1974 | 7.3163 | 6.42E-04 |
| Glucose increasing | EZETIMIBE | 699 | -0.6107 | -1.7336 | 0.5121 | 0.2859 | 145 | 6.4534 | -3.8206 | 16.7275 | 0.2164 |
| Glucose increasing | NORTRIPTYLINE | 2931 | 0.382 | -0.3671 | 1.1311 | 0.3175 | 56 | -7.9286 | -27.2799 | 11.4227 | 0.4151 |
| Glucose increasing | LEVOFLOXACIN | 310 | -1.0102 | -3.3997 | 1.3792 | 0.4061 | 202 | 2.5347 | -6.4778 | 11.5471 | 0.5798 |
| Glucose increasing | CIPROFLOXACIN | 561 | -0.6782 | -2.4179 | 1.0614 | 0.4441 | 142 | 10.5979 | -7.8660 | 29.0618 | 0.2584 |
| Glucose increasing | MOXIFLOXACIN | 77 | -1.3584 | -6.3623 | 3.6456 | 0.5903 | 140 | -0.2695 | -9.1221 | 8.5831 | 0.9521 |
| Glucose increasing | SOTALOL | 586 | 0.5466 | -1.4652 | 2.5584 | 0.5938 | 55 | -2.4909 | -14.8622 | 9.8804 | 0.6880 |
| Glucose increasing | FLUOXETINE | 14162 | -0.0916 | -0.4548 | 0.2717 | 0.6212 | 207 | -1.0700 | -8.6463 | 6.5062 | 0.7809 |
| Glucose increasing | NADOLOL | 291 | 0.6336 | -2.2377 | 3.5049 | 0.6644 | 59 | -9.3393 | -26.8901 | 8.2115 | 0.2912 |
| Glucose increasing | QUETIAPINE | 5131 | 0.1182 | -0.5017 | 0.7381 | 0.7086 | 30 | -13.9333 | -45.7065 | 17.8399 | 0.3772 |
| Glucose increasing | DOXEPIN | 1348 | 0.1244 | -1.1999 | 1.4487 | 0.8538 | 36 | 5.7778 | -14.1608 | 25.7163 | 0.5601 |
| Glucose increasing | SERTRALINE | 29743 | -0.0176 | -0.2644 | 0.2292 | 0.8891 | 248 | 4.1653 | 4.1171 | 12.4477 | 0.3229 |

#### Consistency between glucose and HbA1c findings

Compared with HbA1c, 35 (12.4%) of the 283 medications identified by drug-gene pairs that were included in the glucose analysis. Only one of these medications, oxcarbazepine, an anti-epileptic, had evidence for an effect in both sets. However, the direction of glucose change was inconsistent with glucose increasing in the VA discovery population and decreasing in the VUMC replication. None of the ACE inhibitors demonstrated a consistent impact on glucose in either population. In line with the HbA1c results, verapamil was associated with a consistent decrease in glucose in the VA (−0.82 mg/dL, p=0.03). In the VUMC population, the point estimate indicated that verapamil decreased glucose but the wide confidence interval included the null, potentially due to power limitations of this smaller sample set. There were also inconsistent results for statin medications. In the discovery set, rosuvastatin increased glucose but the remaining statins including simvastatin, atorvastatin, pravastatin, lovastatin, and Fluvastatin, decreased glucose.

#### Meta-analysis

We next performed a cross-site meta-analysis for both HbA1c and glucose in a self-controlled case series to estimate the combined effect for the given medications. The results of these analyses are available in Figures 3 and 4. There was evidence to support all T2D/glucose-reducing medications reducing HbA1c with effect sizes ranging from -0.33% for sitagliptin to -1.06% for glyburide (p<0.001 for all). For glucose, however, only half the T2D/control (−) medications were associated with a mean decrease in glucose. Four (26.7%) of the 15 glucose-increasing medications were associated with an increase in HbA1c as expected, with the most substantial increases being for propranolol and hydrochlorothiazide, respectively (0.12%, p=1.0e^-5^; and 0.08%, p=2.8×10^−14^). Both propranolol and hydrochlorothiazide also demonstrated increases in glucose. Interestingly, there was also some evidence that citalopram and fluoxetine decreased HbA1c (−0.02%, p=0.04; and -0.03%, p=0.01). However, evidence for their impact on glucose was limited. Corticosteroids had the most substantial impact on glucose. Dexamethasone and prednisone consistently increased glucose (7.87 mg/dL, p=9.4×10^−3^; 1.32 mg/dL, p<1.3×10^−16^). In the experimental group, only verapamil demonstrated a significant effect on HbA1c (−0.11%, p=0.01). This effect was also observed in the glucose analysis (−0.85 mg/dL, p=0.024). Three statins, simvastatin, rosuvastatin, and fluvastatin, had evidence for changes in glucose; however, the directions of effect varied among the medications.

**Figure 3.**
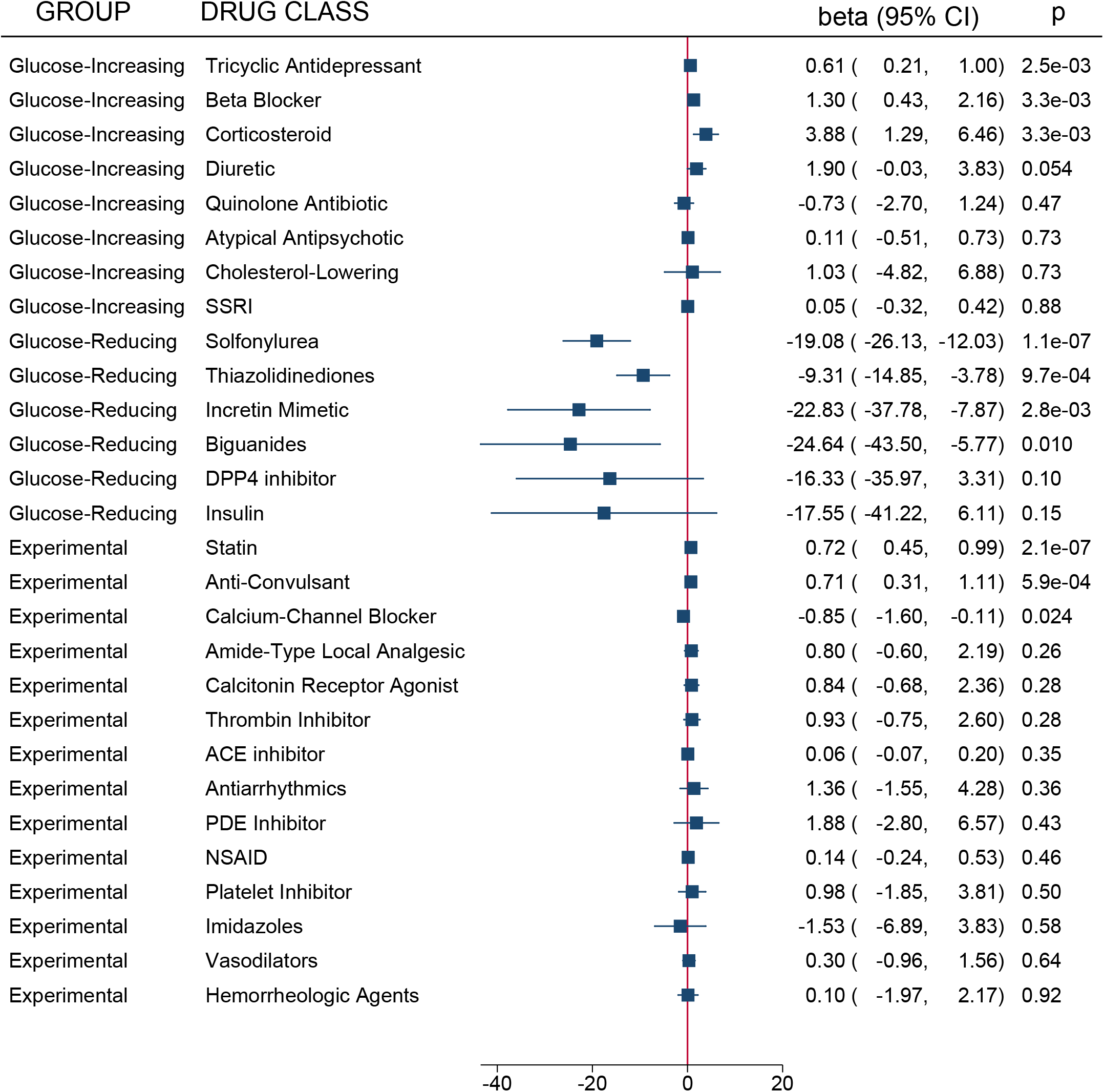
Medication group meta-analysis results from the self-controlled case series for hemoglobin A1c. All medications included in the analysis series were grouped by medication class and meta-analyzed. A forest plot of these results for hemoglobin A1c from the self-controlled case series in the discovery and replication datasets is presented. Whether they belonged to the control, glucose-decreasing or glucose-increasing, or experimental group is noted on the left-hand side of the figure.

**Figure 4.**
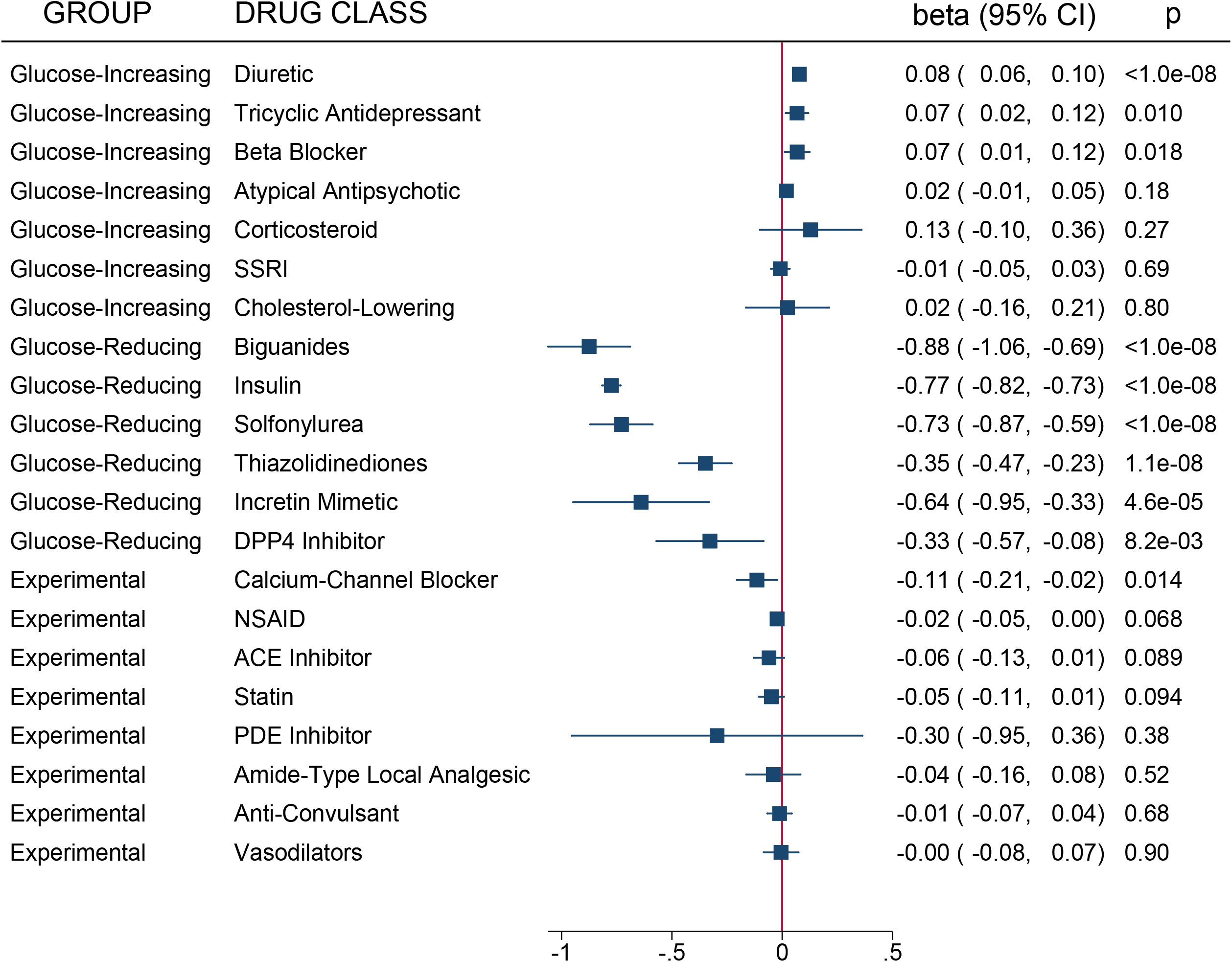
Medication group meta-analysis results from the self-controlled case series for glucose. All medications included in the analysis series were grouped by medication class and meta-analyzed. A forest plot of these results for glucose from the self-controlled case series in the discovery and replication datasets is presented. Whether they belonged to the control, glucose-decreasing or glucose-increasing, or experimental group is noted on the left-hand side of the figure.

To characterize the effect of medication group on HbA1c and glucose, we also performed a drug-class grouped meta-analysis (Figures 3 and 4, respectively). As expected, all six of the T2D/glucose-reducing medication groups lowered HbA1c with the most substantial decrease belonging to biguanides (−0.88%, p<1.6×10^−13^). We saw similar magnitudes of reductions in glucose for DPP4 inhibitors and insulin (p=0.10 and p=0.15, respectively). Three of the glucose-increasing medication groups, diuretics, tricyclic antidepressants, and beta blockers, demonstrated increases in HbA1c and glucose; while corticosteroids only demonstrated an increase in glucose. Only one experimental medication class, calcium channel blockers, substantially reduced HbA1c and glucose (−0.11%, p=0.01; and -0.85 mg/dL, p=0.02, respectively). The statin and anti-convulsant medication classes were associated with increases in glucose (0.72 mg/dL, p=2.1×10^−7^; and 0.71 mg/dL, p=5.9e^-4^, respectively).

### Mendelian randomization

In Table 3, we report our T2D two-sample MR results using S-PrediXcan summary statistics for various indications with putative drug gene targets. In our MR analysis, the strongest evidence for T2D prevention were observed for drugs targeting genes involved in systolic blood pressure, angina, and atrial fibrillation. ACE inhibitors, as proxied by reduced *ACE* gene expression, were predicted to reduce systolic blood pressure by approximately 0.25 mmHg (p=0.28) per standard deviation decrease in *ACE* expression. Whereas for drugs targeting *KCNJ11* expression (i.e., minoxidil and verapamil), the predicted change in systolic blood pressure was minimal per standard deviation change in *KCNJ11* gene expression. A 16-18% T2D risk reduction was observed for ACE inhibitors (T2D OR=0.82, 95% CI=0.78, 0.86, p=3.3×10^−17^) and minoxidil/verapamil (T2D OR=0.84, 95% CI=0.81, 0.87, p=5.0×10^−25^) via changes in predicted systolic blood pressure. Reduced atrial fibrillation risk via decreased expression of *HCN3* and *SCN3A* (via HCN channel blockers and sodium channel blockers, respectively) were associated with reduced T2D risk in the MR analysis; however, only sodium channel blockers were observed to reduce T2D risk (T2D OR=0.25, 95% CI=0.17, 0.39, p=4.7×10^−11^). Similarly, evidence for T2D risk reduction was also observed for angina risk reduction via verapamil as proxied by reduced *CACNA1A* expression (T2D OR=0.17, 95% CI=0.10, 0.29, p=2.1×10^−10^). However, after accounting for potential pleiotropy via changes in correlated indications, only ACE inhibitors demonstrated consistent evidence for a reduction in T2D risk (T2D MVMR OR=0.86, 95% CI=0.84, 0.89, *P*=4.8×10^−06^).

**Table 3.** Summary of estimated Mendelian randomization (MR) effects of drug-targeted genetically-predicted gene expression (GPGE) on indications (either continuous or dichotomous) with type 2 diabetes risk

| Gene | Indication-specific GPGE |  |  |  |  |  |  | T2D MR |  |  |  |  |  |  |
| --- | --- | --- | --- | --- | --- | --- | --- | --- | --- | --- | --- | --- | --- | --- |
| target | Specific drug | Drug effect | No. of | No. of |  |  |  |  |  |  | Egger | MVMR |  |  |
|  | Or drug class | on gene | SNPs<sup>1</sup> | Tissues<sup>1</sup> | Beta<sup>2</sup> | SE | P value | OR<sup>3</sup> | 95% CI | P value | P value<sup>4</sup> | OR<sup>5</sup> | 95% CI | P value |
|  |  | expression |  |  |  |  |  |  |  |  |  |  |  |  |
|  | Systolic blood pressure |  |  |  |  |  |  |  |  |  |  |  |  |  |
| ACE | ACE inhibitors | Antagonist | 108 | 11 | -0.253 | 0.223 | 0.28 | 0.82 | 0.78-0.86 | 3.3x10<sup>-17</sup> | 0.2 | 0.86 | 0.84-0.89 | 4.8x10<sup>-6</sup> |
| KCNJ11 | Minoxidil | Agonist | 454 | 13 | 0.016 | 0.13 | 0.9 | 0.84 | 0.81-0.87 | 5.0x10<sup>-25</sup> | 0.7 | n/a | n/a | n/a |
| KCNJ11 | Verapamil | Antagonist | 454 | 13 | -0.016 | 0.13 | 0.9 | 0.84 | 0.81-0.87 | 5.0x10<sup>-25</sup> | 0.7 | n/a | n/a | n/a |
| TH | Metyrosine | Antagonist | 29 | 2 | 0.061 | 0.323 | 0.88 | 0.79 | 0.43-1.45 | 0.45 | n/a | n/a | n/a | n/a |
| CACNA1A | Verapamil | Antagonist | 1 | 1 | 2.032 | 0.847 | 1.6x10<sup>-2</sup> | 0.64 | 0.56-0.73 | 2.1x10<sup>-10</sup> | n/a | n/a | n/a | n/a |
| GCK | Sitaxentan | Agonist | 66 | 2 | 0.321 | 0.187 | 0.34 | 0.78 | 0.71-0.87 | 2.3x10<sup>-6</sup> | n/a | n/a | n/a | n/a |
|  | Angina |  |  |  |  |  |  |  |  |  |  |  |  |  |
| CACNA1A | Verapamil | Antagonist | 1 | 1 | 0.511 | 0.2 | 1.1x10<sup>-2</sup> | 0.17 | 0.10-0.29 | 2.1x10<sup>-10</sup> | n/a | n/a | n/a | n/a |
|  | Atrial fibrillation |  |  |  |  |  |  |  |  |  |  |  |  |  |
| HCN3 | HCN chan. blockers | Antagonist | 114 | 5 | 0.102 | 0.035 | 4.6x10<sup>-2</sup> | 0.75 | 0.53-1.06 | 0.11 | 1.4x10<sup>-2</sup> | 0.92 | 0.77-1.09 | 0.43 |
| SCN3A | Na+ chan. blockers | Antagonist | 83 | 3 | 0.041 | 0.01 | 5.2x10<sup>-2</sup> | 0.26 | 0.17-0.39 | 4.7x10<sup>-11</sup> | 0.71 | n/a | n/a | n/a |
|  | Bipolar disorder |  |  |  |  |  |  |  |  |  |  |  |  |  |
| SCN3A | Elpetrigine | Antagonist | 46 | 1 | 0.15 | 0.074 | 4.3x10<sup>-2</sup> | 0.92 | 0.69-1.22 | 0.55 | n/a | n/a | n/a | n/a |
|  | Rheumatoid arthritis |  |  |  |  |  |  |  |  |  |  |  |  |  |
| MAPK3 | Sulindac | Antagonist | 95 | 8 | -2.382 | 0.555 | 3.6x10<sup>-3</sup> | 1.01 | 0.99-1.01 | 0.08 | 0.72 | n/a | n/a | n/a |
|  | Glaucoma |  |  |  |  |  |  |  |  |  |  |  |  |  |
| BCL2 | Isosorbide | Antagonist | 46 | 1 | 0.148 | 0.075 | 4.9x10<sup>-2</sup> | 1.01 | 0.85-1.20 | 0.93 | n/a | n/a | n/a | n/a |
|  | Congestive heart failure |  |  |  |  |  |  |  |  |  |  |  |  |  |
| ACE | ACE inhibitors | Antagonist | 28 | 1 | 0.184 | 0.073 | 1.1x10<sup>-2</sup> | 1.31 | 1.02-1.68 | 3.2x10<sup>-2</sup> | n/a | n/a | n/a | n/a |

| Pain |  |  |  |  |  |  |  |  |  |  |  |  |  |  |
| --- | --- | --- | --- | --- | --- | --- | --- | --- | --- | --- | --- | --- | --- | --- |
| *IKBKE* | Amlexanox | Antagonist | 242 | 8 | -0.015 | 0.003 | $6.0 \times 10^{-4}$ | 1.22 | 0.78-1.88 | 0.38 | 0.17 | n/a | n/a | n/a |
| *TAP2* | Olcegepant | Antagonist | 259 | 4 | -0.004 | 0.006 | 0.6 | 0.08 | 0.01-1.36 | 0.08 | 0.82 | n/a | n/a | n/a |
| *OPRL1* | Opioid analgesics | Agonist | 244 | 14 | 0.012 | 0.002 | $1.0 \times 10^{-5}$ | 38.3 | 18.1-81.2 | $1.7 \times 10^{-21}$ | $1.6 \times 10^{-3}$ | n/a | n/a | n/a |
| Coronary artery disease |  |  |  |  |  |  |  |  |  |  |  |  |  |  |
| *HCN3* | Ivabradine | Antagonist | 140 | 6 | -0.076 | 0.028 | $4.0 \times 10^{-2}$ | 1.43 | 1.06-1.93 | $2.0 \times 10^{-2}$ | 0.09 | 1.26 | 1.03-1.54 | $4.9 \times 10^{-2}$ |
| Epilepsy |  |  |  |  |  |  |  |  |  |  |  |  |  |  |
| *SCN3A* | Anticonvulsants | Antagonist | 1 | 1 | 0.058 | 0.028 | $3.7 \times 10^{-2}$ | 0.6 | 0.17-2.08 | 0.42 | n/a | n/a | n/a | n/a |

<sup>1</sup> Number of statistically significant tissues (p value < 0.05) from each indication-specific GPGE model, and the number of SNPs that were used in the GPGE model.
<sup>2</sup> Summarized Indication-specific GPGE effects using random-effects meta-analysis across the statistically significant tissues.
<sup>3</sup> Inverse-variance weighted MR analysis for T2D risk using statistically significant tissues from the GPGE model as instruments.
<sup>4</sup> Egger regression p value < 0.05 indicates the potential for directional pleiotropy.
<sup>5</sup> Multivariable MR (MVMR) used to estimate associations with T2D risk adjusted for GPGE for other correlated indications/traits.
Systolic blood pressure adjusted for congestive heart failure (ACE); atrial fibrillation adjusted for coronary artery disease (HCN3) or bipolar disorder and epilepsy (SCN3A), and coronary artery disease adjusted for atrial fibrillation (HCN3).

## Discussion

Our study has demonstrated an approach for genetics-informed drug repurposing for diabetes by leveraging the power of genomics data, drug annotation databases, EHRs, and contemporary statistical methods for causal inference. A strength of this approach is the rapid and cost-efficient ability to prioritize candidate drugs for clinical trials on the basis of pre-clinical experiments using patient studies and MR.

We demonstrated that calcium-channel blockers are a good target for repurposing for T2D, supported by evidence across all arms of the investigation. This medication demonstrated substantially lower reductions in glycemic indices than the traditional diabetes medications. Our results may be of particular importance to patients with pre-diabetes and hypertension where preferential use of CCBs for blood pressure control may have an additive impact on glycemic control compared with other antihypertensive therapies. We also observed reductions in glycemic indices with another anti-hypertensive medication group, ACE inhibitors. About half of the experimental ACE inhibitors demonstrated significant decreases in both glucose and HbA1c, however, the relative effect sizes varied across sites and depended on the medication type suggesting ACE inhibitor effectiveness to lower glycemic indices depends on type, dose, and possibly the population in which it was used. These findings are similar to clinical trials of ACE inhibitors that have also demonstrated inconsistent results.

The results of this study demonstrate the feasibility and strength of our approach for computational drug repurposing discovery. While many of the potential repurposing medications for glucose reduction have been explored in other trials, we believe these results demonstrate the feasibility of this approach for other disease conditions. Further, because these results derive from two real world clinical populations, they are likely to be robust. These results suggest medications that could be used preferentially to treat concomitant disease while providing potential additional benefit to patients at risk for diabetes or with inadequately controlled diabetes.

This study also demonstrates the importance of careful consideration of study design when performing secondary studies in EHR data sources across sites. The limitations related to use of these resources included ascertainment and utilization of pharmacy data, a continuous measurement that can be compared over time, and the relatively short timeframe under observation. Despite these limitations, the ability to rapidly evaluate impacts on glycemic indices provide a real-world presentation in two distinct cohorts of patients of the effects of the genetically identified medications. The additional support for a causal effect of the identified drugs provided by the MR analyses provides both epidemiological rigor and evidence of a biological mechanism at work in the phenomenon of the glucose-lowering effect of CCBs.

In conclusion, we present a computational approach to drug repurposing that has the potential to be used for other conditions and with other data resources. We demonstrated this approach with a SCCS design in two large EHR systems and with MR using extant results from large-scale GWAS. We identified CCBs as a candidate treatment for high glucose and observed some evidence for ACE inhibitors as well. This strategy can be implemented for other outcomes with access to sufficient quantities of EHR data and informatics expertise.

## Data Availability

Summary level statistical results will be made available upon reasonable request to corresponding authors. Individual level data can be made available following institutional agreements.

## Acknowledgements

The authors wish to acknowledge the efforts of Eric Tortenson and Max Breyer for their assistance with the initial data extraction from Vanderbilt University Medical Center Synthetic Derivative. Figures 1 and 2 were created with BioRender.com.

## Notes

**Conflicts of Interest** No conflicts of interest

**Funding Sources** The dataset used for the replication analyses was obtained from the Vanderbilt University Medical Center Synthetic Derivative, which is supported by institutional funding, the 1S10RR025141-01 instrumentation award, and by the CTSA grant UL1TR000445 from National Center for Advancing Translational Sciences/National Institutes of Health. M.M.S. was supported by AHA 17SFRN33520017 and VA Merit I01 BX005399-01A1. N.K.K. is supported by NIH R00 CA215360. V.W. is supported by the Medical Research Council Integrative Epidemiology Unit at the University of Bristol, UK [MC_UU_00011/4] and the COVID-19 Longitudinal Health and Wellbeing National Core Study, which is funded by the UK Medical Research Council (MC_PC_20059). M.V. and P.R. are supported by 2I01BX003362-03A1. This work was supported using resources and facilities of the US Department of Veterans Affairs (VA), Veterans Health Administration, Cooperative Studies Program, grant number 825-MS-DI-33848, and used resources and facilities at the VA Informatics and Computing Infrastructure (VINCI), VA HSR RES 13-457.

### Competing Interest Statement

The authors have declared no competing interest.

### Funding Statement

The dataset used for the replication analyses was obtained from the Vanderbilt University Medical Center Synthetic Derivative, which is supported by institutional funding, the 1S10RR025141-01 instrumentation award, and by the CTSA grant UL1TR000445 from National Center for Advancing Translational Sciences/National Institutes of Health. M.M.S. was supported by AHA 17SFRN33520017 and VA Merit I01 BX005399-01A1. N.K.K. is supported by NIH R00 CA215360. V.W. is supported by the Medical Research Council Integrative Epidemiology Unit at the University of Bristol, UK [MC_UU_00011/4] and the COVID-19 Longitudinal Health and Wellbeing National Core Study, which is funded by the UK Medical Research Council (MC_PC_20059). M.V. and P.R. are supported by 2I01BX003362-03A1. This work was supported using resources and facilities of the US Department of Veterans Affairs (VA), Veterans Health Administration, Cooperative Studies Program, grant number 825-MS-DI-33848, and used resources and facilities at the VA Informatics and Computing Infrastructure (VINCI), VA HSR RES 13-457.

### Author Declarations

The studies obtained ethical approval from the affiliated organizations, Department of Veterans Affairs and Vanderbilt University Medical Center, Nashville, TN. For the Vanderbilt University Medical Center Synthetic derivative patient data is de-identified and do not meet the 45 CFR 46 definition for human subjects research. As such, the study received non-human subjects determination from the associated institutional review board. For the Department of Veterans Affairs dataset the study was reviewed and approved by institutional review boards and research committees at 3 VA Medical Centers (Salt Lake City, Palo Alto, and West Haven) with approved waivers of informed consent and HIPAA authorization.

